# RAAS blockers and region-specific variations in COVID-19 outcomes: findings from a systematic review and meta-analysis

**DOI:** 10.1101/2020.09.09.20191445

**Authors:** Upinder Kaur, Sankha Shubhra Chakrabarti, Tejas K Patel

## Abstract

**Background:** Coronavirus disease 2019 (COVID-19) has evolved as a global crisis with high mortality seen in elderly and people with cardiometabolic diseases. The use of renin angiotensin aldosterone system (RAAS) blockers in these patients is known to enhance the expression of ACE-2, the chief binding receptor of SARS-CoV-2 and may potentially enhance infectivity.

**Objective:** To provide a pooled estimate of the effect of RAAS blocker usage on COVID-19 outcomes.

**Data Sources:** An electronic literature search was performed for published (using MEDLINE/PubMed and Google Scholar) and preprint (using bioRxiv and medRxiv) studies of interest. The last search was conducted on 9^th^ July 2020.

**Study Selection:** Studies reporting data on RAAS blocker use and COVID-19 mortality and severity were included in the review.

**Data Extraction and Synthesis:** Mortality data and severity data including hospitalization, intensive care unit (ICU) admission, invasive ventilation, steroid use and acute kidney injury (AKI) were recorded. Pooled Odds ratio (OR) estimates were reported with 95% CIs and level of heterogeneity (*I*^2^).

**Main Outcomes and Measures:** Odds of mortality in users of RAAS blockers with respect to non-users was the primary outcome. Odds of severity, hospitalization, ICU admission, mechanical ventilation, steroid use, and AKI in users with respect to non-users of RAAS blockers were the secondary outcomes.

**Results:** Of 1348 articles identified, 48 published studies were included in the final analysis, with a total of 26432 patients from 31 studies included in mortality analysis and 20127 patients from 23 studies included in severity analysis. Majority of the studies (41.6%) were from China. No increased risk of mortality (Pooled OR 0.91 (0.65-1.26), I^2^=89%) or severity (Pooled OR 1.08 (0.79-1.46), I^2^=88%) was seen with RAAS blockers. The drug class was protective in hypertension (pooled OR 0.63 (0.46-0.86), I^2^=58%). Severity of COVID-19 outcomes was found to be high for Europeans (Pooled OR 2.08 (1.52-2.85), I^2^=77%) and US patients (Pooled OR 1.87 (1.62-2.17) in users of RAAS-blockers. A nearly 4 times higher risk of hospitalization, two times higher risk of ICU admission and mechanical ventilation was observed in US patients on RAAS blockers. No net effect on mortality and severity outcomes was seen in Chinese patients. RAAS blocker usage did not have any effect on corticosteroid use and AKI in Chinese patients.

**Conclusions and Relevance:** Use of RAAS blockers is not associated with increased risk of mortality in COVID-19 patients. Reduced mortality is seen in hypertensive patients with COVID-19 and therefore the drugs should be continued in this subset. US and European patients are at higher risk of severe outcomes. Pharmacogenomic differences may explain the ethnicity related variations.

## 1. Introduction

Corona Virus Disease-2019 (COVID-19) caused by Severe Acute Respiratory Syndrome Corona Virus-2 (SARS-CoV-2) has affected 24,652,652 individuals worldwide and claimed 836,065 lives as of 28^th^ August 2020.(1) ACE2 is the major binding receptor of SARS-CoV-2 and is located on pulmonary epithelial cells, endothelial cells and in cells of the kidney, among others. Acute respiratory distress syndrome, myocardial injury, multiorgan failure and disseminated intravascular coagulation (DIC) including diffuse pulmonary intravascular coagulopathy are responsible for majority of the deaths and stem from a state of inflammatory cytokine storm and vascular thrombosis.(2,3) Older individuals and those with co-morbidities such as hypertension, diabetes mellitus (DM) and ischaemic heart disease (IHD) are at increased risk of a severe form of the disease. The use of renin-angiotensin-aldosterone system (RAAS) blockers such as angiotensin converting enzyme inhibitors (ACEIs), angiotensin receptor blockers (ARBs) and mineralocorticoid receptor antagonists (MRAs) in such patients is not uncommon. With experimental evidence of upregulation of ACE2 by RAAS blockers, some concerns were raised related to the increased risk of infection and severity of disease in the users of these drugs.(4,5)

Following this, multiple observational studies were conducted to assess the relationship between the use of RAAS blockers and severity of COVID-19. This systematic review and meta-analysis aim to compile the information obtained from these clinical studies and elucidate the association between the use of RAAS blockers and clinical outcomes in patients of COVID-19. In the past few months, few such meta-analyses have been published but were limited due to the inclusion of small numbers of studies. The current meta-analysis of 48 studies provides the most comprehensive view of the issue till date by involving a much larger number of patients and analysing for multiple health outcomes, as well as by performing region-specific analyses.

## 2. Methods

### 2.1. Search Criteria

A comprehensive search was conducted in PubMed, Google Scholar and the preprint servers medrRxiv.org and bioRxiv.org using keywords: ACEI OR ACE-I OR Angiotensin converting enzyme inhibitors AND COVID-19/SARS-CoV-2, Angiotensin receptor blocker OR AT-1 receptor blocker OR Ang II blocker OR ARB AND COVID-19/SARS-CoV-2, RAAS blocker AND COVID-19/SARS-CoV-2, Aldosterone antagonist AND COVID-19/SARS-CoV-2, Renin inhibitor AND COVID-19/SARS-CoV-2. The final search was conducted on 9^th^ July 2020. Only articles published in English language were included in this study.

### 2.2. Selection Criteria

#### 2.2.1. Inclusion criteria

- All clinical studies (observational studies and clinical trials) analysing the effect of RAAS blockers on clinical outcomes in patients of laboratory confirmed COVID-19 were included in this study. Thus, the review involved inclusion of studies which compared the disease outcomes between users and non-users of RAAS blockers as well as those which assessed the use of RAAS blockers in COVID-19 patients of varying severity. The term RAAS blockers include ACEIs, ARBs, aldosterone antagonists and renin inhibitors. Studies were considered irrespective of the dose and duration of RAAS blocker use.
- Studies should have provided comparative data of mortality and/or severity between users and non-users of RAAS blockers in patients of COVID-19.
- All types of study setting (outpatient, inpatient, nursing homes, home care approach) were included.
- All age groups of study population were included

#### 2.2.2. Exclusion criteria

- Studies focusing on individual RAAS blockers only.
- Studies focusing only on outcomes based on laboratory parameters (e.g., serum or urinary ACE2 expression).
- Non-comparative studies, review articles, in-vitro studies, animal studies, viewpoints.

All relevant abstracts were scrutinized, and full text was searched for those found useful. In case of lack of clarity in the abstracts, full text was analysed. This was done by author UK assisted by author SSC and confirmed by author TKP assisted by author SSC.

### 2.3. Data Extraction

From the included studies, data was extracted in a Microsoft Excel Sheet. Data included author name, publication year, country, study design, total duration of study, mean or median follow up, characteristics of patients or specific population of COVID-19 patients in whom the particular study was conducted, age, gender, sample size, use of RAAS blockers, mortality outcomes, severity outcomes, need of hospitalization, care in intensive care unit (ICU), need of mechanical ventilation, corticosteroid use and occurrence of acute kidney injury (AKI).

### 2.4. Risk of bias

Two investigators (TKP and SSC) assessed the risk of bias in the included studies as per the Newcastle - Ottawa quality assessment scale (NOS) adapted for the cross-sectional design. The criteria considered were representativeness of the study sample, sample size, non-respondents, ascertainment of the exposure, comparability of study groups for the confounders (age and major co-morbidities), assessment of outcome and statistical tests. The maximum possible score was 10.(6)

### 2.5. Outcomes

The primary outcome was odds of mortality in the users of RAAS blockers with respect to non-users among confirmed cases of COVID-19. The secondary outcomes were odds of severity, hospitalization, ICU admission, mechanical ventilation, steroid use, and AKI in users of RAAS blockers with respect to non-users. In the absence of universally accepted definitions, severity was considered as defined by the authors in the included studies. When outcomes were reported both under ‘critical’ and ‘severe’ headings, we considered the more serious outcome under severity analysis. In case of multiple time-point for the outcome estimation, we considered data at the end of study period.

A subgroup analysis of all outcomes was performed based on the geographical locations (country or continent of origin) of the included studies. The mortality outcome was further analysed as per study sub-populations (e.g., patients with hypertension). The severity outcome was stratified based on definitions considered by individual authors. A sensitivity analysis was performed for each outcome after excluding studies with high risk of bias. The studies with score ≤ 7 on the modified NOS scale were considered to have high risk of bias.

### 2.6. Data synthesis

All outcomes were dichotomous variables. They are reported as odds ratio (OR) with 95% confidence intervals (CI). The meta-analysis was weighted with inverse variance method. An I^2^ test was used to assess the heterogeneity between the studies. Fixed-effect model was used if heterogeneity was less than 50% and random-effect was applied in case heterogeneity exceeded 50%. Funnel plot method was used for reporting publication bias. The meta-analysis was performed using Review Manager Software version 5.4.

## 3. Results

### 3.1. Characteristics of included studies

As shown in **Figure 1**, a total of 1348 articles were retrieved. Out of 70 full text articles assessed, 48 studies satisfying the selection criteria were included for detailed qualitative and quantitative analysis in this review. **Table 1** shows the demographic features of the patients in included studies.(3,7–52) Majority of the studies assessing the outcomes of interest have been reported from China (N=20, 41.6%) followed by Europe (N=16, 33.3%). The sample size of individual studies varied from 36 to 9519. In case of 31 (64.5%) studies, mean or median age of patients was more than 60 years.

**Figure 1:**
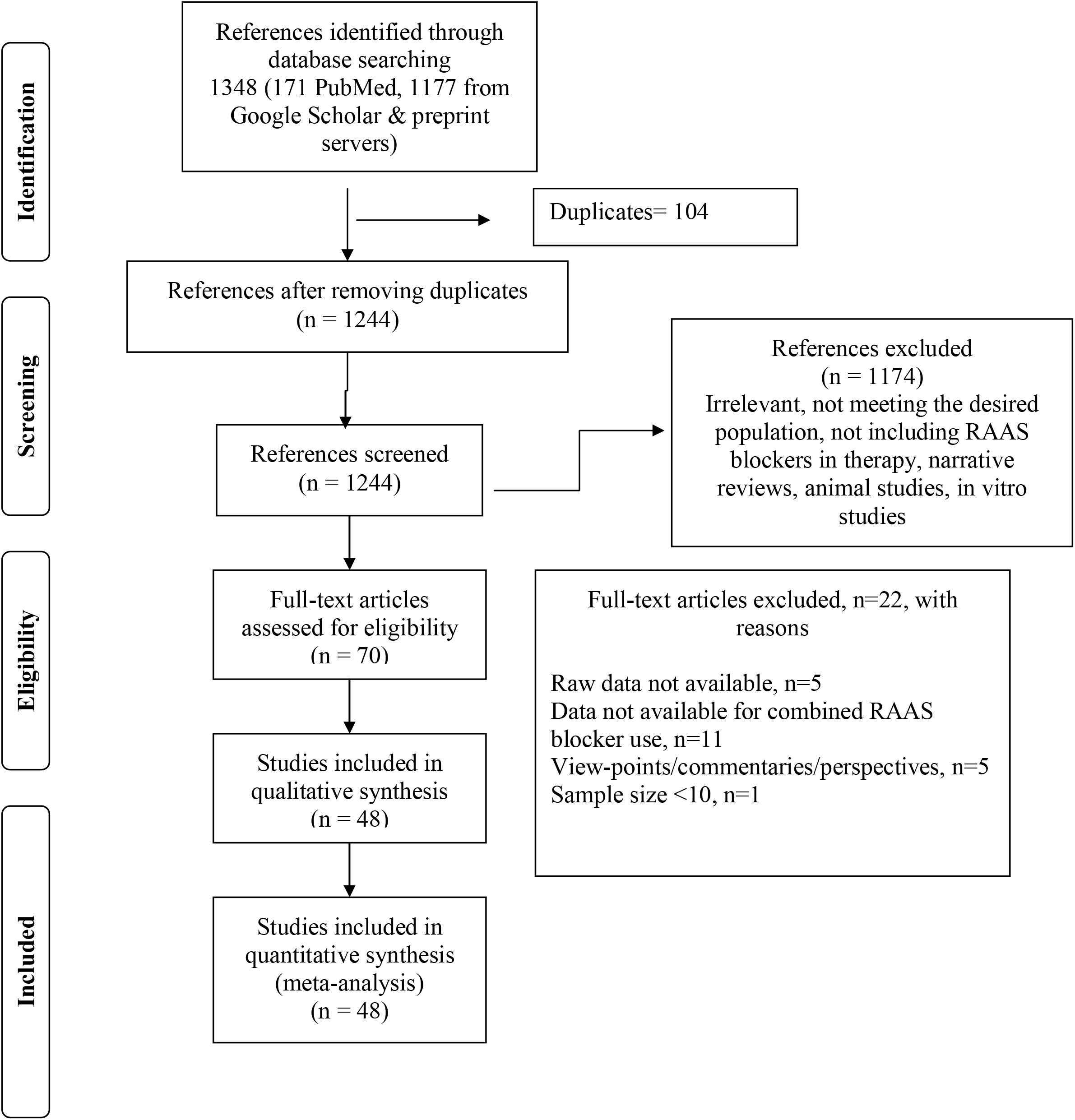
Flow diagram of number of studies screened and selected

**Table 1.** Studies included in meta-analysis of impact of renin angiotensin aldosterone system blockers on mortality and severity outcomes of COVID-19.

| Author | Country | Design | Total duration | Follow up per patient | Age (in years) | Sample size studied (M/F) | Indication for RAAS blocker | Outcome tested | Co-morbidities | Confounder adjustment (For included variables) | Quality (NOS score) |
| --- | --- | --- | --- | --- | --- | --- | --- | --- | --- | --- | --- |
| Andrea C | Italy | Retrospective | 43d | 28d | Mean (SD) entire sample 63.4 (14.9) | 191 (131/60); HTN=96 | HTN | Mortality | In HTN gp-CKD: 41.6%, CAD: 28.1%, DM: 22.9% | No | 8 |
| Argenziano MG | USA | Retrospective | 61d | NM | Median (IQR) entire sample 63 (50-75) | 1000 (596/404) | NM | Severity assessed as hospitalization/ICU admission/IV | HTN: 60%, DM: 37%, CAD: 13% | No | 7 |
| Baker KF | UK | Retrospective | 119d | 28d | Median (IQR) 75 (60-83) | 316 (173/143) | NM | Mortality | HTN: 42%, Respiratory diseases: 32%, DM: 26.6% | Yes | 10 |
| Bean DM | UK | NM | 22d | 7d | Mean (SD) entire sample 63 (20) | 205 (106/99) | NM | Severity ¶ | HTN: 51%, DM: 30%, IHD/HF: 14.6% | Yes | 10 |
| Bravi F | Italy | Retrospective | NM | 24d | Mean (SD) entire sample 58 (20.9) | 1603 (758/845); HTN=543 | HTN | Severity ¶ | HTN: 34%, Major CVD: 16%, DM: 12% | Yes | 10 |
| Caraballo C | USA | Retrospective | NM | NM | Median entire sample (IQR) 78 (65-87) | 206 (93/113) | HF | Mortality | HTN: 80%, Renal disease: 38.3%, CAD: 35.4% | No | 8 |
| Chen Y | China | Retrospective | 77d | NM | Median entire sample (IQR) 58 (42-62) | 71 (with known history of medication) | DM & HTN | Mortality | HTN: 36.6%, CVD: 14.7%, DM: 14.4% | No | 6 |
| Choi HK | South Korea | Retrospective | 116d | NM | Mean (SD) Users gp- 65 (13)<br>Non-users group- 68 (15) | 1585 (679/906) | HTN | Mortality & Severity $\phi^a$ | Users gp: DM: 46.5%, Major neurologic diseases: 28%<br>Non-users gp: DM: 43%, Major neurologic diseases: 42.7% | Yes | 9 |
| de Abajo F | Spain | Case Control | 24d | NM | Mean (SD) entire sample 69.1 (15.4) | 1139 (695/444) | NM | Severity ¶ | HTN: 54.2%, DLP: 39%, DM: 272% | Yes | 10 |
| Du B | China | Retrospective | 40d | NM | Median (IQR) Users gp- 71 (63.5-77)<br>Non-users gp- 69 (62-77) | 154 (79/75) | Raised Troponin I | Mortality | Users gp- HTN: 100%, DM: 41.2%, CVD: 29.4%<br>Non-users gp- HTN: 38.7%, DM: 19%, CVD: 18.3% | Yes | 7 |
| Dublin S | USA | Retrospective | 106d | NM | Mean (SD) entire tested sample 66 (12.2) | 56105 (29455/26650); Tested positive = 720 | NM | Severity assessed as hospitalization | Users gp- HTN: 71.5%, DM: 33.5%, Renal disease: 12.2%<br>Non-users gp- | Yes | 10 |
|  |  |  |  |  |  |  |  |  | HTN: 9.9%,<br>DM: 3.8%,<br>Renal<br>disease:<br>2.3% |  |  |
| Felice C | Italy | Retrospective | 23d | NM | Mean<br>(SD)<br>ACE<br>users<br>73.1<br>(11.5)<br>ARB<br>users<br>69<br>(13.4)<br>Non-users<br>76.2<br>(11.9) | 133 (86/47) | HTN | Mortality<br>&<br>Severity<br>assessed<br>as<br>H/ICU/<br>Non-IV | Users gp-DM: 24%,<br>Cancer:<br>17%,<br>COPD: 9%<br>Non-users<br>gp-DM: 28%,<br>Cancer:<br>14%,<br>COPD: 14<br>% | Yes | 9 |
| Feng Y | China | Retrospective | 46d | NM | Median<br>(IQR)<br>entire<br>sample<br>53 (40-64) | 476<br>(271/205) | NM | Severity<br>* | DM: 10.3%,<br>CVD: 8%,<br>COPD:<br>4.6% | No | 4 |
| Feng Z | China | Retrospective | 59d | NM | Median<br>(IQR)<br>entire<br>sample<br>47 (36-58) | 564<br>(284/280) | HTN | Severity<br>* | HTN:<br>14.5%,<br>DM: 8%,<br>CVD: 3.9% | Yes | 10 |
| Fosbol EL | Denmark | Retrospective | 94d<br>(73d of<br>Nested CC) | NM | Median<br>(IQR)<br>entire<br>sample<br>54.7<br>(40.9-72)<br>Users<br>gp-72.8<br>(61.0-81.0) | 4480<br>(2144/<br>2336) | NM | Mortality<br>&<br>Severity<sup>\$</sup> | Users gp-HTN:<br>70.8%,<br>DM: 24.2%,<br>MI: 21.6%<br>Non-users<br>gp- HTN:<br>5.8%,<br>DM: 5.4%,<br>MI 5.2% | Yes | 10 |
|  |  |  |  |  | Non-users<br>gp-50.1<br>(37.2-64.5) |  |  |  |  |  |  |
| Gao C | China | Retrospective | 57d | Median<br>(IQR)<br>21d<br>(12d-32d) | Mean<br>(SD)<br>64.24<br>(11.2) | 850<br>(443/407) | HTN | Mortality<br>&<br>Severity* | Users gp-DM: 30.1%,<br>Angina:<br>17.5%,<br>PCI/CABG:<br>4.9%<br>Non-users<br>gp-DM: 26.6%,<br>Angina:<br>15.2\$,<br>PCI/CABG:<br>5.3% | Yes | 10 |
| Golpe R | Spain | Retrospective | 24d | Mean<br>(SD)<br>22d<br>(7d) | Mean<br>(SD)<br>70.4<br>(12.3) | 157 (72/85) | HTN | Severity<br>assessed<br>as<br>hospitaliz<br>ation | DM: 33.7%,<br>DLP: 51.6% | No | 9 |
| Guo T | China | Retrospective | 32d | NM | Mean<br>(SD) of<br>entire<br>sample<br>58.5<br>(14.66) | 187 (91/96) | NM | Mortality | HTN:<br>32.6%,<br>DM: 15%,<br>CHD:<br>11.2% | No | 8 |
| Hu J | China | NM | 23d | NM | Median<br>(IQR)<br>57<br>(49.5-66) | 149 (88/61) | HTN | Mortality<br>&<br>Severity* | Users gp-DM: 24.6%,<br>CLD: 7.7%,<br>Renal<br>disease:<br>6.1%<br>Non-users<br>gp- DM:<br>16.7%,<br>CLD:<br>4.76%,<br>Renal<br>disease:<br>2.4% | No | 5 |
| Huang L | China | Retrospective | 40d | NM | Mean (SD) entire sample 58 (17) | 200 (115/85) | NM | Mortality & Severity assessed by OF/IV<sup>a</sup> | Users gp-HTN: 75%, DM: 25%, CHD: 18%<br>Non-users HTN: 22%, DM: 14%, CHD: 8% | No | 8 |
| Huang Z | China | Retrospective | 26d | NM | Mean (SD)<br>Users gp- 52.65 (13.12)<br>Non - users gp- 67.77 (12.84) | 50 (27/23) | HTN | Mortality & Severity* | Users gp-COPD, Anaemia: 5%, CAD, DM: 0%<br>Non-users gp-DM: 13.3%, CAD: 3.3%, COPD, anaemia: 0% | No | 6 |
| Inciardi RM | Italy | NM | 22d | 14d minimum | Mean (SD) entire sample 67 (12) | 99 (80/19) | NM | Mortality | HTN: 64%, DM: 31%, DLP: 30% | No | 6 |
| Ip A | USA | Retrospective | NM | NM | <50 to >80 years | 1584 with HTN, 1216 with known outcomes | HTN | Mortality | NM | No | 7 |
| Jung SY | South Korea | Cohort study | NM | NM | Mean (SD)<br>Users gp- 62.5 (14.7)<br>Non-users gp- | 5179 (2295/2884) | NM | Mortality & Severity assessed as IV | Users gp-HTN: 94%, DM: 48%, COPD: 40%<br>Non-users gp-COPD: 27%, DM: 11%, HTN: 10%, | Yes | 10 |
|  |  |  |  |  | 41.5<br>(16.6) |  |  |  |  |  |  |
| Li J | China | Retrospective | 61d | NM | Median<br>(IQR)<br>entire<br>sample<br>55.5<br>(38-67)<br>HTN<br>cohort<br>66 (59-73) | 362<br>(189/173) | HTN | Mortality<br>&<br>Severity* | Users gp-DM: 36.5%,<br>CbVD,<br>CHD:<br>23.5%<br>Non-users<br>gp-DM: 34.4%,<br>CHD:<br>14.2%,<br>CbVD:<br>16.6% | No | 7 |
| Li X | China | Retrospective | 38d | 32d | Median<br>(IQR)<br>entire<br>sample<br>60 (48-69) | 548<br>(279/269) | NM | Severity# | HTN:<br>30.3%,<br>DM: 15.1%,<br>CHD: 6.2% | No | 8 |
| Liabeuf S | France | Retrospective | 47d | NM | Median<br>(IQR)<br>73 (61-84) | 268<br>(164 on at<br>least one<br>anti HTN) | NM | Mortality<br>&<br>Severity<br>assessed<br>as ICU<br>admission | HTN: 57%,<br>type 2 DM:<br>18%,<br>Stroke: 14% | Yes | 10 |
| Mehta N | USA | Retrospective | 36d | NM | Mean<br>(SD)<br>ACEI<br>gp 63<br>(15)<br>ARB<br>gp 65<br>(13) | 1735 | NM | Mortality<br>&<br>severity<br>assessed<br>as<br>hospitaliz<br>ation/<br>ICU<br>admission/<br>IV | ACEI users<br>gp vs non -<br>users gp-HTN: 97%<br>vs 37%,<br>DM: 54%<br>vs 17%,<br>CAD: 21%<br>vs 9%<br>ARB users<br>gp vs non-users gp-HTN: 93%<br>vs 38%,<br>DM: 50%<br>vs 18%,<br>CAD: 24 %<br>vs 9% | Yes | 10 |
| Meng J | China | Retrospective | 44d | NM | Median (IQR)<br>64.5 (55.8-69) | 42 (24/18) | HTN | Mortality & Severity* | Users gp-DM & CHD: 29.4%<br>Non-users gp- DM & CHD: 32% | No | 6 |
| Mohamed MMB | Australia | NM | NM | Median 25d, minimum 14d | Non-AKI gp- 66 (23-97)<br>AKI gp- 65 (34-96) | 575 (312/263) | NM | Severity assessed as AKI | HTN: 73.7%,<br>DM: 48.8% | No | 7 |
| Otero DL | Spain | Retrospective | 28d | NM | Mean (SD)<br>59.5 (20.3)<br>Users gp-72.1 (13.2)<br>Non-users gp- 56 (20.5) | 965 (425/540) | NM | Mortality & Severity assessed as hospitalization/HF/ICU admission & composite of HF/death | Users gp-HTN: 98.6%,<br>DLP: 60%,<br>DM: 27.6%<br>Non-users gp-DLP: 19.3%,<br>HTN: 12.1%,<br>DM: 8.7% | Yes | 10 |
| Oussalah A | France | Retrospective | 31d | NM | Median (IQR)<br>65 (54-77) | 149 (91/58) | NM | Mortality & Severity assessed as acute respiratory failure/IV | Users gp-HTN: 86%,<br>DM: 58%,<br>CVD: 49%<br>Non-users gp-HTN: 32%,<br>CVD: 19%,<br>DM: 14% | Yes | 10 |
| Regina J | Switzerland | Retrospective | 25d | 14d minimum | Median (IQR)<br>70 (55-81) | 200 (120/80) | NM | Severity assessed as IV | HTN: 43.5%,<br>DM: 21.5%,<br>CAD: 17.5% | No | 8 |
| Reilev M | Denmark | NM | 64d | 30d | Median (IQR) entire sample | 9519 (4010/5509) | NM | Mortality & Severity assessed | HTN: 25%,<br>Chronic lung | No | 8 |
|  |  |  |  |  | 49 (34-63) |  |  | as ICU admission/hospitalization | disease: 13%, IHD: 9.1% |  |  |
| Rentsch CT | USA | Retrospective | 52d | NM | Median (IQR) 66.1 (60.4-71) | 585 (558/27) | NM | Severity assessed as hospitalization/ICU admission | HTN: 72.3%, DM: 44.4%, Vascular disease: 27.9% | Yes | 10 |
| Reynolds HR | USA | NM | 46d | NM | Median (IQR) full sample 49 (34-63)<br>HTN sample 64 (54-75) | 5894; HTN=2573 | HTN | Severity ¶ | HTN: 34.6%, DM: 18%, OLD: 14.6% | Yes | 10 |
| Rhee SY | South Korea | NM | NM | NM | Mean (SD) Users gp- 64.85 (13.23)<br>Non-users gp- 59.9 (16.7) | 832 (445/387) | DM | Severity ¶ | Users gp-HTN: 99.7%, DLP: 92.6, CVD: 31.2%<br>Non-users gp-HTN: 47.9%, DLP: 84.5%, CVD: 24.7% | Yes | 9 |
| Richardson S | USA | NM | 35d | NM | Median entire sample (IQR) 63 (52-75) | 5700 (3437/226; HTN with known outcomes= 1366 | HTN | Mortality & Severity assessed as ICU admission/IV/AKI | HTN: 56.6%, DM: 33.8&, CAD 11.1% | No | 6 |
| De Spiegeleer A | Belgium | Retrospective | 47d | NM | Mean (SD)<br>86 (7) | 154 (51/103) | NM | Severity<br>ϕϕ | Users gp- HTN: 93.3%, DM: 20%<br>Non-users gp- HTN: 8.8%, DM: 17.8% | Yes | 9 |
| Tan ND | China | Retrospective | 71d | 30d | Median (IQR)<br>67 (62-70) in users gp,<br>67.5 (57-71) in non-users gp | 100 (?51/49) | HTN | Mortality & Severity* | Users gp- DM: 25.8%, GI illness: 19.4%, CHD: 16.1%<br>Non-users gp- DM: 29%, GI illness: 24.6%, CHD: 18.8% | No | 5 |
| Trecarichi EM | Italy | Retrospective | 41d | NM | Mean (SD)<br>80 (12) | 50 (24/26) | NM | Mortality | CVD: 82%, Neurologic disease: 52%, Psychiatric disease: 30% | No | 6 |
| Xie Y | China | Retrospective | 32d | NM | <65 years- 36.3%<br>≥65 years- 63.7% | 102 (46/56) | ACRI | Mortality & Severity* | HTN: 54%, DM: 22.5%, CHD: 14.7% | No | 7 |
| Xu J | China | Retrospective | 59d | NM | Median (IQR)<br>65 (58-73) | 101 (53/48) | HTN | Mortality & Severity<br>¶ | Users gp- HTN: 100%, DM: 20%, CHD: 13%<br>Non-users gp- HTN: 100%, | Yes | 10 |
|  |  |  |  |  |  |  |  |  | DM: 18%,<br>CHD: 11% |  |  |
| Yang G | China | Retrospective | 59d | NM | Median (IQR)<br>66 (61-73),<br>Users gp- 65 (57-72)<br>Non-users gp- 67 (62-75) | 126 (62/64) | HTN | Mortality & Severity* | Users gp-DM: 30.2%, Cardiopathy : 16.3%, Neurologic disease: 9.3%<br>Non-users gp-DM: 30%, Cardiopathy : 19.3%, Neurologic disease: 7.2% | No | 8 |
| Zeng Z | China | Retrospective | 64d | 14d minimum | Mean (SD)<br>HTN gp- 67 (11)<br>Users gp- 64 (12)<br>Non-users gp- 69 (10) | 75 (41/34) | HTN | Mortality & Severity# | DM: 31%, CVD: 21%, CbVD: 15% | No | 6 |
| Zhang P | China | Retrospective | 68d | 28d | Median (IQR)<br>Users gp- 64 (55-68),<br>Non-users gp- 64 (57-69) | 1128 (603/525) | HTN | Mortality (as HR, raw data not available) & Severity assessed as IV & steroid use | Users gp-DM: 23.4%, CHD: 15.4%, CRD: 3.7%<br>Non-users gp- DM: 20.9%, CHD: 10.9%, CRD: 3% | No | 7 |
| Zhou J | China | Retrospective | 145d | NM | Median (IQR)<br>35 (32-37) | 1043 (563/480),<br>n=976 with known medication history | NM | Severity assessed as ICU admission | Respiratory diseases: 43%,<br>GIT diseases: 32.5%,<br>HTN: 20.2% | No | 10 |
| Zhou X | China | Retrospective | 27d | NM | Mean (SD)<br>57.7 (14.2)<br>HTN gp-64.8 (10.1) | 110 (60/50),<br>n=36 with HTN (19/17) | HTN | Mortality | HTN: 32.7%, DM: 10%,<br>CVD: 9.1% | Yes | 6 |

[‘Users’ refers to patients on RAAS blockers, ‘non-users’ refers to those not on RAAS blockers
¶: Severity in terms of composite of ICU or CCU admission/death; ϕ: Severity in terms of composite of death/ severe infection, latter including respiratory failure or organ failure leading to mechanical ventilation, ICU admission, RRT and ECMO; \*: Severity as per Severity Criteria of National Health Commission of China; \$: Severity as per SARS/ ICU admission; #: Severity as defined by American Thoracic Society and Infectious Diseases Society of America; !: Valsartan Sacubitril was also taken as ARB; ?: Study did not mention male-female distribution clearly; ϕϕ: DOH ≥ 7d or death; a: The criteria resembled ‘Critical’ of Chinese criteria and the study data was therefore analysed under the subgroup of ‘critical’ outcomes as per Chinese definition
Abbreviations: Major CVD: major cardiovascular disease (congestive heart failure, myocardial infarction, or stroke); ACRI: Acute cardiac related injury; AKI: Acute kidney injury; CVD: Cardiovascular disease; CbVD: Cerebrovascular disease; CAD: Coronary artery disease; CHD: Coronary Heart Disease; COPD: Chronic obstructive pulmonary disease; D: Death; DLP: Dyslipidaemia, DM: Diabetes mellitus; d: days; ECMO: Extra corporeal membrane oxygenation; GIT: gastrointestinal tract; gp: group; HF: heart failure; HTN: hypertension; ICU: Intensive Care Unit; IHD: ischemic heart disease; IV: invasive ventilation; NM: not mentioned; NOS: Newcastle-Ottawa scale; OF: organ failure; OLD: Obstructive lung disease; RAAS: Renin angiotensin aldosterone system; RRT: Renal replacement therapy]

A total of 32 (66.7%) studies assessed mortality out of which 31 were included in mortality analysis as raw data was not available in the study by Zhang P et al.(45) A total of 35 studies (77.8%) assessed composite severity or individual health outcomes (hospitalization, ICU admission, mechanical ventilation, steroid use, and AKI). Twelve studies defined severity as per clinical guidelines of the National Health Commission of China. Six studies defined severity as the composite of ICU admission and death. Two studies used the severity definition issued by the Infectious Diseases Society of America (IDSA). Composite of hospitalization for ≥ 7 days and death, composite of death/ severe infection (definition described in the table legend), and composite of SARS/ ICU admission were taken as severe outcome in one study each. In seventeen studies, severity was considered based on individual health outcomes such as ICU admission, invasive ventilation, AKI, and hospitalization as mentioned in **Table 1**. RAAS blockers were used for hypertension in 21 studies (43.7%) while indication of their use was not mentioned in 25 studies (52%). Duration of follow up was mentioned in 14 studies and ranged from 7-32 days. Confounder adjustment had been performed in 22 (45.8%) studies for two major confounders. A total of 30 studies were considered to have low risk of bias.

### 3.2. Mortality analysis

A total of 26432 patients from 31 studies (6030 users of RAAS blockers and 20402 non-users) were included in the mortality analysis. The use of RAAS blockers was not associated with increased risk of mortality (pooled OR 0.91 (0.65-1.26), I^2^= 89%) (**Figure 2**). A similar trend was observed in the sensitivity analysis after excluding studies with high risk of bias (pooled OR 1.09 (0.71-1.67, I^2^= 91%). Funnel plot was asymmetrical on visual inspection (**Supplementary Figure 1**).

**Figure 2.**
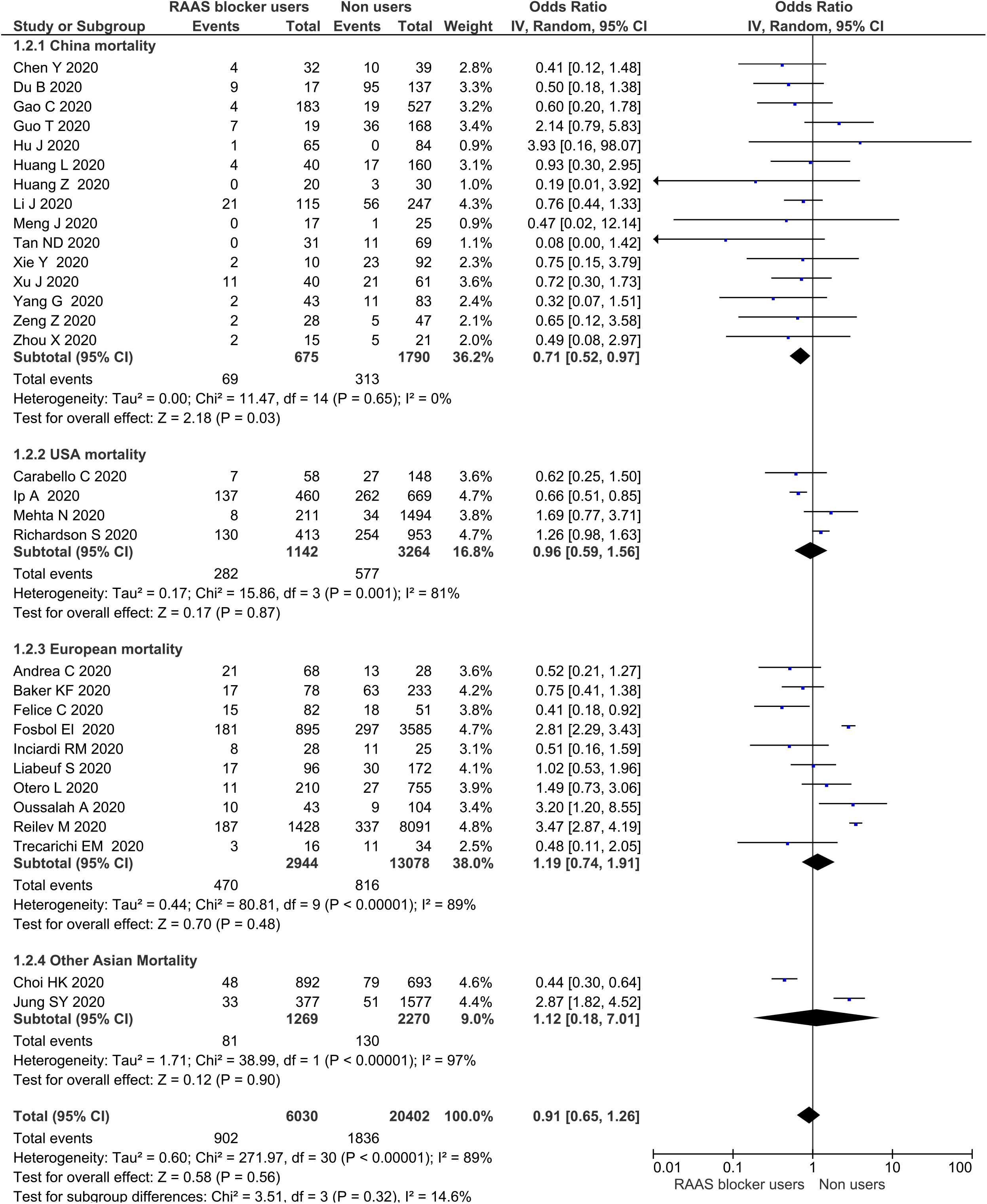
Overall and region-specific mortality effects of RAAS blockers in COVID-19 patients

The subgroup analysis of mortality outcome based on geographical locations showed use of RAAS blockers conferred a protection from mortality in the Chinese population (OR 0.71 (0.52-0.97)) (**Figure 2**). However, in sensitivity analysis, no difference in mortality was observed in studies with low risk of bias (pooled OR 0.85 (0.48-1.50), I^2^ =25%). Neither benefit nor risk was observed with the use of RAAS blockers in patients in the US (pooled OR 0.96 (0.59-1.56), I^2^ = 81%), Europe (pooled OR 1.19 (0.74-1.91), I^2^ =89%), and South Korea (pooled OR 1.12 (0.18-7.01), I^2^ =97%) (**Figure 2**). The results were consistent in sensitivity analysis (**Table 2**).

**Table 2.** Meta-analysis of all outcomes-summary of results.

| **Parameter** | **Number of studies (number of patients)** | **OR (CI)** | **OR (CI) Sensitivity analysis** |
| --- | --- | --- | --- |
| **Mortality** | 31 (26432) | 0.89 (0.64-1.24), $I^2=89\%$ | 1.09 (0.71-1.67), $I^2=91\%$ |
| **Country/Region specific mortality** |  |  |  |
| China | 15(2465) | 0.71 (0.52-0.97), $I^2=0\%$ | 0.85 (0.48-1.50), $I^2=25\%$ |
| Europe | 10 (16022) | 1.19 (0.74-1.91), $I^2=89\%$ | 1.37 (0.84-2.23), $I^2=90\%$ |
| USA | 4 (4406) | 0.96 (0.59-1.56), $I^2=81\%$ | 1.04 (0.39-2.81), $I^2=64\%$ |
| Other Asian (South Korean) | 2 (3539) | 1.12 (0.18-7.01), $I^2=97\%$ | 1.12 (0.18-7.01), $I^2=97\%$ |
| **Severity** | 23 (20127) | 1.08 (0.79-1.46), $I^2=88\%$ | 1.32 (0.93-1.87), $I^2=91\%$ |
| **Definition wise severity** |  |  |  |
| ‘Critical’ (Chinese classification) | 8 (3396) | 0.50 (0.33-0.76), $I^2=29\%$ | 0.63 (0.28-1.45), $I^2=70\%$ |
| ‘Severe’ (Chinese classification) | 4 (571) | 0.71 (0.30-1.69), $I^2=54\%$ | 0.14 (0.02-1.13) |
| ICU/death composite | 6 (9941) | 1.82 (1.31-2.53), $I^2=82\%$ | 1.82 (1.31-2.53), $I^2=82\%$ |
| Severity (IDSA/ATS) | 2 (620) | 1.36 (0.49-3.80), $I^2=69\%$ | 0.86 (0.45-1.61) |
| Others | 3 (5599) | 2.14 (1.22-3.74), $I^2=69\%$ | 2.14 (1.22-3.74), $I^2=69\%$ |
| **Country/Region wise severity** |  |  |  |
| China | 13 (3002) | 0.69 (0.45-1.06), I<sup>2</sup>=51% | 0.68 (0.3-1.53), I<sup>2</sup>=58% |
| Europe | 7 (8814) | 2.08 (1.52-2.85), I<sup>2</sup>=77% | 2.08 (1.52-2.85), I<sup>2</sup>=77% |
| USA | 1 (5894) | 1.87 (1.62-2.17) | 1.87 (1.62-2.17) |
| Other Asians | 2 (2417) | 0.62 (0.32-1.23), I<sup>2</sup>=69% | 0.62 (0.32-1.23), I<sup>2</sup>=69% |
| **Disease wise mortality** |  |  |  |
| HTN | 15 (6060) | 0.63 (0.46-0.86), I<sup>2</sup>=58% | 0.48 (0.36-0.63), I<sup>2</sup>=0% |
| Not specified | 12 (19839) | 1.58 (1.1-2.27), I<sup>2</sup>=82% | 1.81 (1.28-2.58), I<sup>2</sup>=81% |
| Others<sup>#</sup> | 4 (533) | 0.55 (0.31-0.96), I<sup>2</sup>=0% | 0.62 (0.25-1.50) |
| **Hospitalisation** | 7 (13849) | 2.1 (1.09-4.05), I<sup>2</sup>=96% | 2.36 (1.2-4.65), I<sup>2</sup>=95% |
| **Country/Region wise hospitalisation** |  |  |  |
| USA | 4 (4040) | 2.86 (1.13-7.24), I<sup>2</sup>=97% | 3.87 (1.21-12.34), I<sup>2</sup>=97% |
| Europe | 3 (9809) | 1.17 (0.20-6.82), I<sup>2</sup>=95% | 1.17 (0.20-6.82), I<sup>2</sup>=95% |
| **ICU admission** | 13 (16441) | 1.37 (0.86-2.19), I<sup>2</sup>=91% | 1.55 (0.79-3.02), I<sup>2</sup>=93% |
| **Country/Region wise ICU admission** |  |  |  |
| USA | 4 (3376) | 1.47 (1.15-1.87), I<sup>2</sup>=37% | 1.82 (1.29-2.58), I<sup>2</sup>=0% |
| Europe | 4 (10154) | 1.51 (0.57-4.03), I<sup>2</sup>=93% | 1.51 (0.57-4.03), I<sup>2</sup>=93% |
| China | 3 (350) | 0.67 (0.35-1.27), I<sup>2</sup>=0% | 0.65 (0.25-1.68), I<sup>2</sup>=0% |
| Other Asians | 2 (2561) | 2.64 (0.08-85.87), I<sup>2</sup>=97% | 2.64 (0.08-85.87), I<sup>2</sup>=97% |
| **Invasive ventilation** | 15 (10678) | 1.06 (0.7-1.59), I<sup>2</sup>=80% | 1.28 (0.58, 2.83), I<sup>2</sup>=88% |
| **Invasive ventilation country wise** |  |  |  |
| USA | 3 (4101) | 2.33 (1.02-5.36), I<sup>2</sup>=92% | 9.72 (4.35-21.71) |
| Europe | 2 (446) | 0.64 (0.17-2.46), I<sup>2</sup>=86% | 0.64 (0.17-2.46), I<sup>2</sup>=86% |
| China | 8 (2592) | 0.79 (0.55-1.14), I<sup>2</sup>=0% | 1.03 (0.45-2.37), I<sup>2</sup>=50% |
| Other Asians | 2 (3539) | 1.24 (0.27-5.66), I<sup>2</sup>=92% | 1.24 (0.27-5.66), I<sup>2</sup>=92% |
| **Corticosteroid use** | 7 (1854) [All from China] | 0.82 (0.65-1.04), I<sup>2</sup>=38% | 1.01 (0.64-1.6), I<sup>2</sup>=35% |
| **AKI** | 5 (2143) | 0.94 (0.76-1.16), I<sup>2</sup>=0% | 1.23 (0.52-2.89), I<sup>2</sup>=0%<br>(Based on 2 Chinese studies) |

[<sup>#</sup>One study each of patients with heart failure, acute cardiac related injury, diabetes mellitus and hypertension, and elevated cardiac biomarkers
*Abbreviations:* AKI: Acute kidney injury, ATS: American Thoracic Society, CI: Confidence Interval, IDSA: Infectious Disease Society of America, OR: Odds Ratio, ICU: Intensive Care Unit]

On indication or disease-wise comparison, use of RAAS blockers was found to reduce the overall risk of mortality when prescribed for hypertension (pooled OR 0.63 (0.46-0.86), I^2^=58%). Similar trend was observed in sensitivity analysis (pooled OR 0.48 (0.36-0.63), I^2^=0%). Ten out of fifteen studies reporting mortality in hypertensive patients were from China. (**Figure 3**). The results were consistent in sensitivity analysis (pooled OR 1.81 (1.28-2.58), I^2^ =81%).

**Figure 3.**
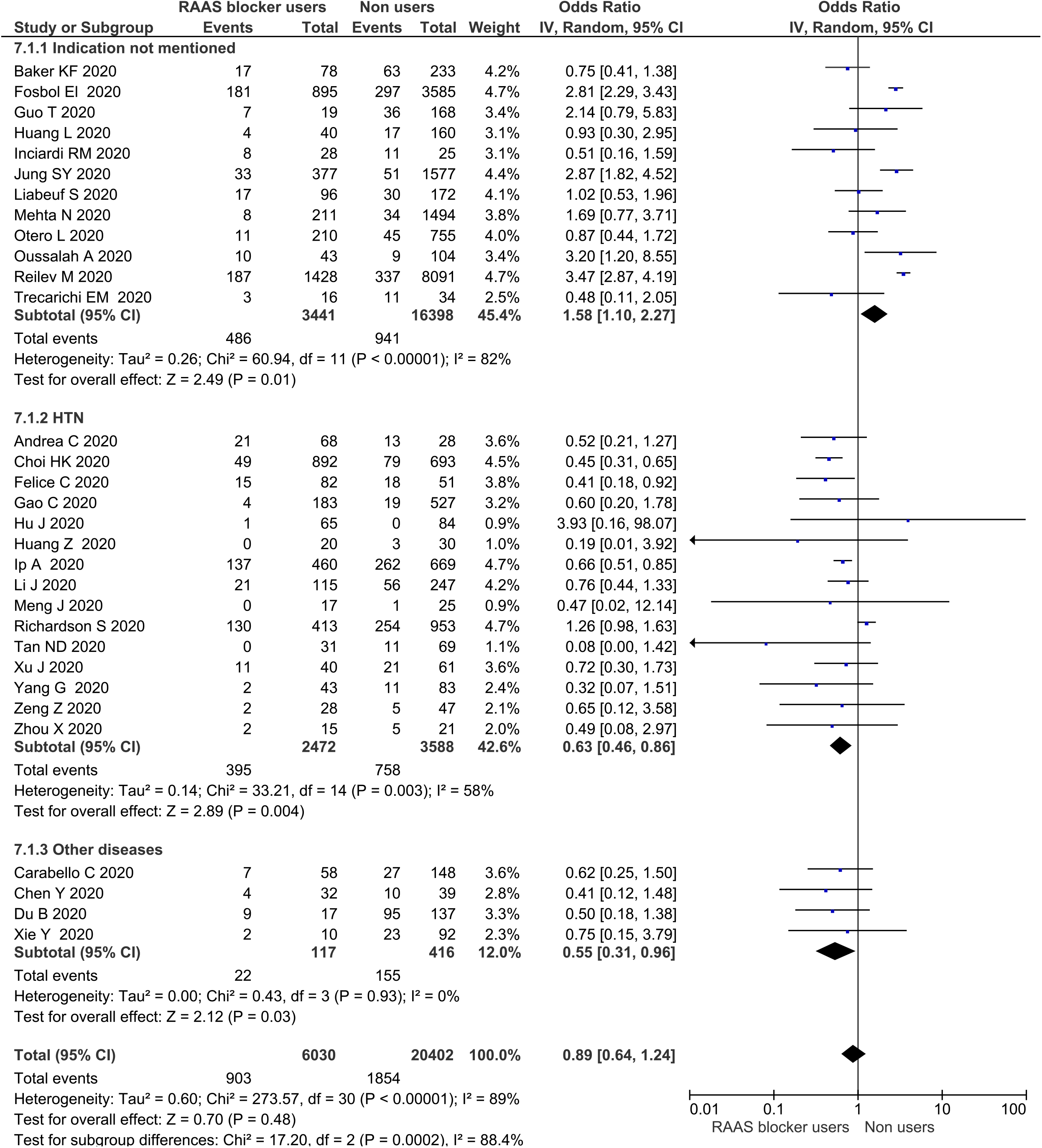
Disease/indication specific mortality effects of RAAS blockers in COVID-19 patients

### 3.3. Severity analysis

A total of 20127 patients (5460 RAAS blocker users & 14667 non-users) from 23 studies were included in the severity analysis. The overall pooled summary showed no effect on the severity of disease with the use of RAAS blockers (pooled OR 1.08 (0.79-1.46), I^2^=88%) (**Figure 4**). A similar result was observed in sensitivity analysis (pooled OR 1.32 (0.93-1.87), I^2^=91%). Funnel plot was asymmetrical on visual inspection (**Supplementary Figure 2**).

**Figure 4.**
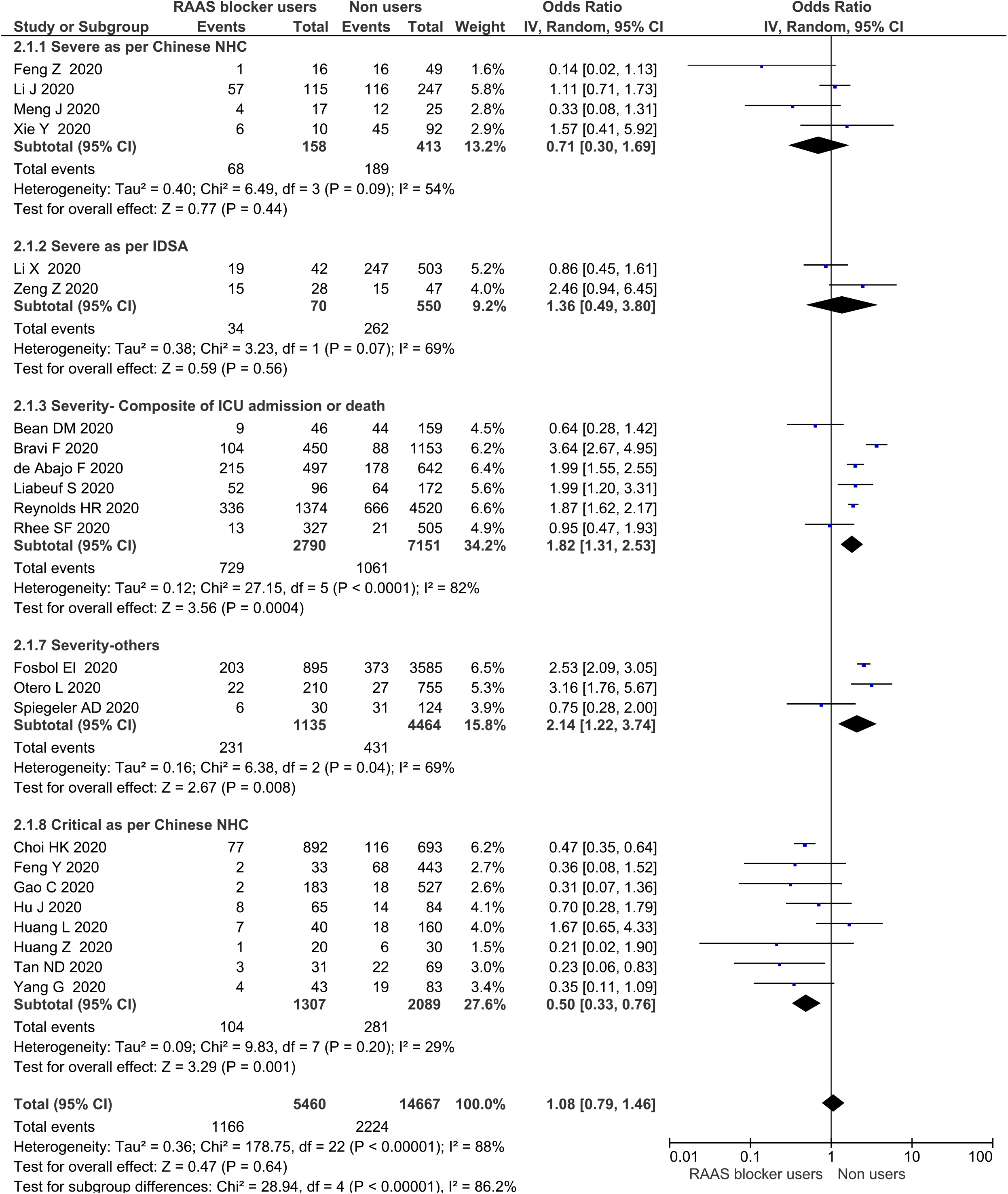
Effects of RAAS blockers on severity of COVID-19 disease (pooled and definition-specific)

Comparison of studies with respect to the definition of severity showed a protective effect of RAAS blockers against ‘critical’ disease defined by National Health Commission of China (pooled OR 0.5 (0.33-0.76), I^2^=29%). Seven out of eight studies assessing this parameter were from China. The effect, however, was nullified on sensitivity analysis (pooled OR 0.63 (0.28-1.45), I^2^=70%). On the other hand, RAAS blockers were found to increase the risk of composite outcome of ICU and death (pooled OR 1.82 (1.31-2.53), I^2^=82%) with a similar trend in sensitivity analysis. Among the four studies showing negative impact of RAAS blockers, three involved the European population, one enrolled US patients while none was from China (**Figure 4**).(24,34,40,51)

Region/country specific analysis also showed an increased risk of poor health outcomes in European patients (pooled OR 2.08 (1.52-2.85), I^2^=77%) and US patients (OR 1.87 (1.62-2.17)) (**Figure 5**). A similar trend was observed in sensitivity analysis. In contrast, no effect on severity with the use of RAAS blockers was evident in the Chinese population in overall (pooled OR 0.69 (0.45-1.06), I^2^=51%), and sensitivity analysis (pooled OR 0.68 (0.3-1.53), I^2^=58%) (**Figure 5**).

**Figure 5.**
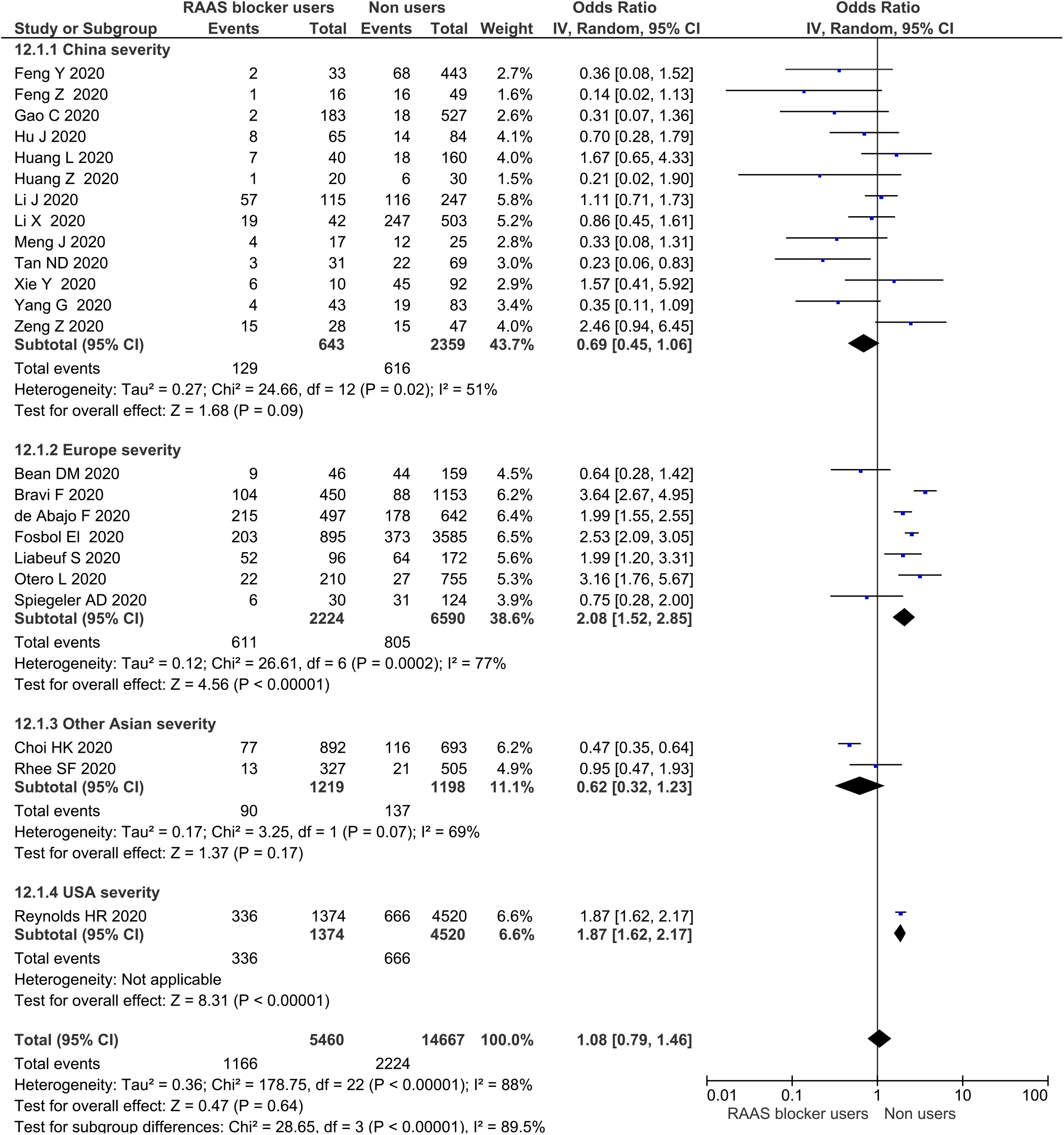
Region-specific severity effects of RAAS blockers in COVID-19 patients

### 3.4. Hospitalization

Risk of hospitalization was analysed in seven studies with 13849 patients (2565 RAAS blocker users and 11284 non-users). The use of RAAS blockers was associated with increased risk of hospitalization in overall analysis (pooled OR 2.1 (1.09-4.05), I^2^=96%) as well as in sensitivity analysis (pooled OR 2.36 (1.2-4.65), I^2^=95%). Among the seven studies, four involved US patients, three enrolled Europeans while none was from China.(8,9,14,18,25,32,33) Country specific subgroup and sensitivity analysis showed a nearly 4 times higher risk of hospitalisation in US patients (pooled OR 3.87 (1.21-12.34), I^2^=97%) while no such risk was evident in Europeans (pooled OR 1.17 (0.20-6.82), I^2^=95%) (**Supplementary Figure 3**).

### 3.5. ICU admission

A total of 16441 patients (4060 RAAS blocker users and 12381 non-users) from 13 studies were analysed for the assessment of risk of ICU admission. No increased risk of ICU admission was observed with the use of RAAS blockers in the overall (pooled OR 1.37 (0.86-2.19), I^2^=91%) and sensitivity analyses (pooled OR 1.55 (0.79-3.02), I^2^=93%). Country-specific analysis showed an increased risk of ICU admission in the US population in overall (pooled OR 1.47 (1.15-1.87), I^2^= 37%) and sensitivity analyses (pooled OR 1.82 (1.29-2.58), I^2^=0%). No effect on ICU admission was observed in Chinese patients (pooled OR 0.65 (0.25-1.68), I^2^=0%) or in Europeans (pooled OR 1.51 (0.57-4.03), I^2^=93%) (**Supplementary Figure 4**).

### 3.6. Invasive ventilation

Need for invasive ventilation was assessed in 15 studies with a total of 10318 patients. Use of RAAS blockers was not associated with increased requirement of invasive ventilation (pooled OR 1.06 (0.7-1.59), I^2^=80%) and the result did not vary in sensitivity analysis (pooled OR 1.28 (0.58-2.83), I^2^=88%). Country-specific analysis showed an increased risk of invasive ventilation in the US population (pooled OR 2.33 (1.02-5.36), I^2^=92%). After excluding the studies with a high risk of bias, sensitivity analysis could be performed on one study by Mehta et al which showed a significantly high risk of invasive ventilation with RAAS blocker usage (OR 9.72 (4.35-21.71)).(25) No such risk was seen in the Chinese population (pooled OR 0.79 (0.55-1.14), I^2^=0%) or in Europeans (pooled OR 0.64 (0.17-2.46), I^2^=86%). Similar trends were observed in the Chinese and Europeans in sensitivity analysis (pooled OR for Chinese population 1.03 (0.45-2.37), I^2^= 50%; pooled OR for Europeans 0.64 (0.17-2.46), I^2^=86%) (**Supplementary Figure 5**).

### 3.7. Corticosteroid use

Seven studies (n=1854) commented on corticosteroid use in relation to RAAS blocker use. All of these were from China. Use of RAAS blockers did not affect the requirement for corticosteroid use in the overall analysis (pooled OR 0.82 (0.65-1.04), I^2^=38%) and also in the sensitivity analysis (pooled OR 1.01 (0.64-1.6), I^2^= 35%) (**Supplementary Figure 6**).

### 3.8. Acute kidney injury

Five studies (n=2143) reporting on AKI were analysed. Use of RAAS blockers was not associated with increased or decreased risk of AKI in overall analysis (pooled OR 0.94 (0.76-1.16), I^2^=0%) and also in the sensitivity analysis (pooled OR 1.23 (0.52-2.89), I^2^=0%). The latter was based on two studies, both from China (**Supplementary Figure 7**).(16,53)

## 4. Discussion

Hypertension, diabetes mellitus, cerebrovascular disease, and ischemic heart disease are co-morbidities which are commonly prevalent and found to be responsible for adverse prognosis in patients with COVID-19.(54) RAAS blockers are used in majority of these diseases and are known for their disease modifying roles in ischemic heart disease, congestive heart failure, and diabetic nephropathy. With the observation that SARS-CoV-2 binds preferentially to ACE2 as its receptor and that ACE2 is prone to upregulation by RAAS blockers, speculations were made that the continuation of RAAS blockers would increase binding of the virus to host cells and enhance its infectivity. On the contrary, ACE2 is known to be protective against lung injury via the Ang (1-7)-Mas-Mrg D axis.(55,56) Ang (1-7) exerts cardiopulmonary protection via vasodilatory, anti -inflammatory, anti-thrombotic and anti-hypertrophic roles.(57) Downregulation of ACE2 has been shown to exaggerate the lung injury and decrease the overall survival of mice subjected to agents with potential pulmonary toxicity.(55,56) Some clinical studies and pooled analyses have shown a protective role of ACEIs against pneumonia particularly in elderly patients with hypertension and diabetes mellitus.(58,59) Considering this, some groups have hypothesized that upregulation of ACE2 by RAAS blockers might be protective once the virus has entered host cells and therapies causing enhancement of ACE2 might be useful tools in the COVID-19 armamentarium.(60) The confusion surrounding the use of RAAS blockers led to a spurt of observational studies with a focus on establishing relationship if any, between the use of RAAS blockers and COVID-19 outcomes. In this systematic review, we have tried to compile information from all such studies and provide insights on association between the use of RAAS blockers and COVID-19 morbidity and mortality outcomes.

In our meta-analysis, use of RAAS blockers was not associated with an increased risk of mortality. A reduced risk of mortality was seen in the Chinese population, but the effect was nullified in sensitivity analysis. RAAS blockers were found to reduce mortality in hypertensive patients. On the other hand, an increased risk of composite outcome of ICU admission/death was seen with the use of RAAS blockers and this effect persisted in sensitivity analysis.

With respect to severity of COVID-19 disease, although no overall effect of RAAS blockers was evident, a reduced risk of ‘critical’ form of the disease (defined as per NHC China) was observed. The same protection was not validated, however, in sensitivity analysis. Further, while RAAS blockers did not produce any adverse effect on disease severity when analysed in the entire population, the outcomes differed considerably between the countries. RAAS blockers were found not to affect disease severity in Chinese patients but the use of such agents was associated with nearly a two times higher risk of severe disease in US patients and Europeans. Nearly a four times increased risk of hospitalisation was seen with the use of RAAS blockers in US patients. Similarly, no increase in the risk of ICU admission and invasive ventilation was seen with RAAS blockers in Chinese patients, whereas the US patients on RAAS blockers had an approximately two times higher risk of getting admitted in the ICU or receiving mechanical ventilation. Further, with respect to requirement of corticosteroids and causation of renal injury, no risk could be attributed to RAAS blockers. This interpretation is primarily based on the sensitivity analysis involving Chinese studies.

These country specific variations could be due to the interplay of genetic factors which may include but are not limited to polymorphisms involving *ACE* or *ACE2* genes. *ACE2* gene is prone to multiple polymorphisms. Traditionally, *ACE2* polymorphisms have been associated with hypertension as well as reduced blood pressure lowering response to ACEIs.(61) Some of the polymorphisms seen predominantly in Europeans such as K26R can enhance the interaction between S protein of SARS-CoV-2 and ACE2 which might lead to increased severity of the disease.(62) A preprint analysed the relationship between *ACE2* polymorphisms and severity of COVID-19 disease in a small cohort of 62 patients. Notably, single nucleotide polymorphisms (SNPs) increasing the tissue expression of ACE2 were associated with higher rates of hospitalization while a lower odds of severe disease was seen with SNPs decreasing the tissue expression of ACE2.(63) *ACE* I/D genotype can also influence the severity of COVID-19 pneumonia. Polymorphisms involving *ACE* can influence the circulating and tissue levels of ACE as well as of cytokines like IL-6 and kallikreins. Higher enzyme and cytokine levels are seen in those with ID and DD genotypes.(64) *ACE* DD genotype has been shown to be associated with increased cardiovascular morbidity and increased risk of pneumonia in some studies.(65,66) The pneumonia protective potential of ACEIs is commonly observed in Asians and is linked with *ACE* II and ID genotypes prevalent in the Asian population.(67,68)

A recently published study assessed the relationship between allele frequency ratio of *ACE* I/D genotype and COVID-19 recovery. A trend of lesser severity and early recovery was observed with increasing I/D allele ratio. The study showed that I/D ratio of > 1 is seen in China, Japan and East Asia which are some of the less severely affected countries. On the other hand, I/D ratio of less than 1 (0.4-0.6) has been observed for countries like Italy, the US, Spain, Brazil, and the UK, which are affected the most by the COVID-19 pandemic.(69) The sole contribution of genotypic variations behind severity and mortality is however unlikely as some countries like India have an I/D ratio of around 0.11 but have considerably low mortality and severity rates of COVID-19 compared to the West. Environmental, biological and immunological factors can also have additive or decisive roles in modulating COVID-19 severity and mortality.(70,71)

The neutral effect of RAAS blockers on mortality and a protective effect in hypertensives, are consistent with the results of some of the already published meta-analyses. However, among these, the study by Pranata specifically included COVID-19 patients with hypertension while those by Grover and Zhang included a major study by Mehra et al which has now been retracted.(72–74) The number of studies included in these systematic reviews varied from 12 to 16; moreover, severity definition varied considerably across the studies and therefore was difficult to interpret. By incorporating a much larger number of studies in our meta-analysis, we could analyse the correlation between RAAS blocker use and severity as per various definitions. Finally, the review tends to associate the use of RAAS blockers with multiple outcomes such as need for hospitalization, ICU admission, invasive ventilation, steroid use and renal insult, which as per our knowledge, have not been addressed in any pooled analyses so far.

This systematic review has some limitations. The pooled analysis is mainly based on observational studies, which are more likely to have study populations with difference in baseline characteristics and co-interventions than randomized controlled trials. The country specific subgroup analysis was based on only a small number of studies. Further, the current meta-analysis aimed to generate data related to RAAS blockers and therefore excluded those studies (n=11) which focussed on ACEI and ARB class in isolation and did not provide information about the outcomes in combined RAAS blocker class. We did not compare the outcomes between users of ACEIs and ARBs also. However, such analyses can be done in the future to deduce any class specific differences that can potentially influence COVID-19 outcomes.

## 5. Conclusion

There is a need to investigate racial or region/country specific differences in the clinical outcomes of COVID-19. Genetic polymorphisms may govern the pharmacodynamic response to RAAS blockers in different population groups, as seen in our meta-analysis and should be explored actively in future. There is a need to explore excess risk of ICU admission and mechanical ventilation in the US and increased severity of COVID-19 disease in Europeans, both of which were found to be associated with RAAS blocker usage. Overall, the use of RAAS blockers does not seem to have any impact on COVID-19 mortality and severity. In the presence of a protective effect in patients with hypertension, it may be advisable to continue these drugs in those patients with pre-COVID indication for the same.

## Data Availability

Data available in public domain.

## Acknowledgement

Dr Patel and Dr Kaur wish to thank the administration of the All India Institute of Medical Sciences, Gorakhpur for providing research support.

